# Generic probabilistic modelling and non-homogeneity issues for the UK epidemic of COVID-19

**DOI:** 10.1101/2020.04.04.20053462

**Authors:** Anatoly Zhigljavsky, Roger Whitaker, Ivan Fesenko, Kobi Kremnizer, Jack Noonan, Paul Harper, Jonathan Gillard, Thomas Woolley, Daniel Gartner, Jasmine Grimsley, Edilson de Arruda, Val Fedorov, Tom Crick

## Abstract

Coronavirus COVID-19 spreads through the population mostly based on social contact. To gauge the potential for widespread contagion, to cope with associated uncertainty and to inform its mitigation, more accurate and robust modelling is centrally important for policy making.

We provide a flexible modelling approach that increases the accuracy with which insights can be made. We use this to analyse different scenarios relevant to the COVID-19 situation in the UK. We present a stochastic model that captures the inherently probabilistic nature of contagion between population members. The computational nature of our model means that spatial constraints (e.g., communities and regions), the susceptibility of different age groups and other factors such as medical pre-histories can be incorporated with ease. We analyse different possible scenarios of the COVID-19 situation in the UK. Our model is robust to small changes in the parameters and is flexible in being able to deal with different scenarios.

This approach goes beyond the convention of representing the spread of an epidemic through a fixed cycle of susceptibility, infection and recovery (SIR). It is important to emphasise that standard SIR-type models, unlike our model, are not flexible enough and are also not stochastic and hence should be used with extreme caution. Our model allows both heterogeneity and inherent uncertainty to be incorporated. Due to the scarcity of verified data, we draw insights by calibrating our model using parameters from other relevant sources, including agreement on average (mean field) with parameters in SIR-based models.

We use the model to assess parameter sensitivity for a number of key variables that characterise the COVID-19 epidemic. We also test several control parameters with respect to their influence on the severity of the outbreak. Our analysis shows that due to inclusion of spatial heterogeneity in the population and the asynchronous timing of the epidemic across different areas, the severity of the epidemic might be lower than expected from other models.

We find that one of the most crucial control parameters that may significantly reduce the severity of the epidemic is the degree of separation of vulnerable people and people aged 70 years and over, but note also that isolation of other groups has an effect on the severity of the epidemic. It is important to remember that models are there to advise and not to replace reality, and that any action should be coordinated and approved by public health experts with experience in dealing with epidemics.

The computational approach makes it possible for further extensive scenario-based analysis to be undertaken. This and a comprehensive study of sensitivity of the model to different parameters defining COVID-19 and its development will be the subject of our forthcoming paper. In that paper, we shall also extend the model where we will consider different probabilistic scenarios for infected people with mild and severe cases.

## 1 Introduction

We model the development of the COVID-19 epidemic in the UK under different scenarios of handling the epidemic. There are many standard epidemiological models for modelling epidemics, see e.g. [1, 2]. In this research, we use a more generic simulation model which has the following features:

- it can take into account spatial heterogeneity of the population and heterogeneity of development of epidemic in different areas;
- it allows the use of time-dependent strategies for analysing the epidemics;
- it allows taking into account special characteristics of particular groups of people, especially people with specific medical pre-histories and elderly.

Standard epidemiological models such as SIR and many of its modifications do not possess these properties and hence are not applicable for any study that requires the use of the features above. In particular, In particular, for influenza the mortality changes less significantly with age in comparison to coronavirus; hence common influenza models do not give much insight in modelling the COVID-19 epidemic. The report [3], which is widely considered as the main document specifying the current epidemic in the UK and in the world, is almost entirely based on the use of standard epidemiological models and hence the conclusions of [3] seem to be lacking specifics related to important issues of the study such as heterogeneity of development of epidemic at different locations and even flexible use of different death rates across different ages. Moreover, as noted in [4], the basic assumptions for the model of [3] were written more than 13 years ago and based on the specific dynamics of a flu pandemic and hence the model was not calibrated for COVID-19. Unreliability of COVID-19 data, including the numbers of COVID cases and COVID deaths, is a serious problem. It is discussed, in particular, by G. Antes, in [5].

Despite the SIR models including the model of [3] have been heavily criticised [4, 6], when choosing the main parameters of our model we calibrate it so that its mean field version approximately reproduces the same output as SIR-based models of [3, 6] and we use the parameter values consistent with [3, 6]. Also, the main notation (which sometimes does not look natural from a statistician’s point of view) is taken from [6].

The primary objective of our work is construction of a reliable, robust and interpretable model describing the epidemic under different control regimes. In this paper, we make the first step towards this objective. Due to lack of time and unreliability of available data on COVID-19, most of our results serve as an illustration of the role of different control options and we hope that even as illustration they are useful. However, we try to stay close to the COVID-19 epidemic scenario and hence we use appropriate recommendations about the choice of parameters and models of the virus behaviour we found from the studies based on the use of standard models.

See conclusions in Sections 3,4,5 for the main conclusions of this work.

## 2 The model

### Variables and parameters

- *t* - time (in days)
- *t*_0_ - intervention time (e.g., the time when self-isolation starts)
- *N* - population size
- *I*(*t*) - number of infected at time *t*
- *R*(*t*) - number of immune (recovered or dead) at time *t* (this makes full sense from a modelling perspective, but we appreciate that this may look kind of strange when read by non-scientists)
- *S*(*t*) = *N* − *I*(*t*) − *R*(*t*) - number of susceptible at time *t*
- *M* - number of groups selected for the study
- *N*_*m*_ - size of *m*-th group (*N* = *N*_1_ + *N*_2_ + … + *N*_*M*_)
- *I*_*m*_(*t*) - number of infected at time *t* in group *m*
- *R*_*m*_(*t*) - number of immune at time *t* in group *m*
- *S*_*m*_(*t*) - number of susceptible at time *t* in group *m*
- *β* - average number of transmissions of the virus per unit of time with no intervention
- 1*/σ* - average infectious period
- *k* - shape parameter of the Erlang distribution
- *R*_0_ = *β/σ* - reproductive number (average number of people who will capture the disease from one contagious person)
- 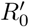- reproductive number after intervention
- *p*_*m*_ - probability to recover for an infected person from the *m*-th group

### Values of parameters and generic model

The reproductive number *R*_0_ is the main parameter defining the speed of development of an epidemic. There is no true value for *R*_0_ as it varies in different parts of the UK (and the world). In particular, in rural areas one would expect a considerably lower value of *R*_0_ than in London. Authors of [3] suggest *R*_0_ = 2.2 and *R*_0_ = 2.4 as typical; the authors of [6] use values for *R*_0_ in the range [2.25, 2.75]. We shall use the value *R*_0_ = 2.5 as typical which may be a slightly pessimistic choice overall but could be an adequate choice for the mega-cities where the epidemics develop faster and may lead to more causalities. In rural areas, in small towns, and everywhere else where social contacts are less intense, the epidemic is milder.

We assume that the person becomes infected *τ* days after catching the virus, where *τ* has Poisson distribution with mean of 1 week. To model the time to recover (or die) we use *Erlang distribution* with shape parameter *k* = 3 and rate parameter *λ* = 1*/*7 so that the mean of the distribution is *k/λ* = 1*/σ* = 21 (in simulations, we discretise the numbers to their nearest integers). This implies that we assume that the average longevity of the period of time while the infected person is contagious is 21 days, in line with the current knowledge, see e.g. [7, 8, 9]. Standard deviation of the chosen Erlang distribution is approximately 12, which is rather large and reflects the uncertainty we currently have about the period of time a person needs to recover (or die) from COVID-19. An increase in 1*/σ* would prolong the epidemic and smaller values of 1*/σ* would make it to cause people to be contagious for less time. The use of Erlang distribution is standard for modelling similar events in reliability and queuing theories, which have much in common with epidemiology. We have considered the sensitivity of the model in this study with respect to the choice of parameters *λ* and *σ* but more has to be done in cooperation with epidemiologists. As there are currently many outbreaks epidemics, new knowledge about the distribution of the period of infection by COVID-19 can emerge soon.

The model varies depending on purposes of the study. The main ingredients are: *I*_*m*_(*t*) is the birth-and-death processes and *R*_*m*_(*t*) is the associated pure birth processes. The process of transmission is the Poisson process with intensity *β* (time to next transmission has the exponential density *βe*^−*βt*^, *t >* 0). After the intervention (for *t* ≥ *t*_0_), the Poisson process of transmission for *m*-the group has intensity *β*_*m*_. We treat this model as purely stochastic despite parts of it can be written it terms of systems of stochastic differential equations. Despite running pure simulation models taking longer than running combined models, they are simpler and less prone to certain errors.

### The number of infected at time *t* as the main quantity of interest

To start with, in Figure 1 we consider an uninterrupted run of an epidemic with *R*_0_ = 2.5 in a homogeneous (one-group) population. The starting time of an epidemic is unknown and is even hard to define as the first transmissions of the virus take random and perhaps long times. We start plotting the curves after 0.5% of the population are infected.

**Figure 1:**
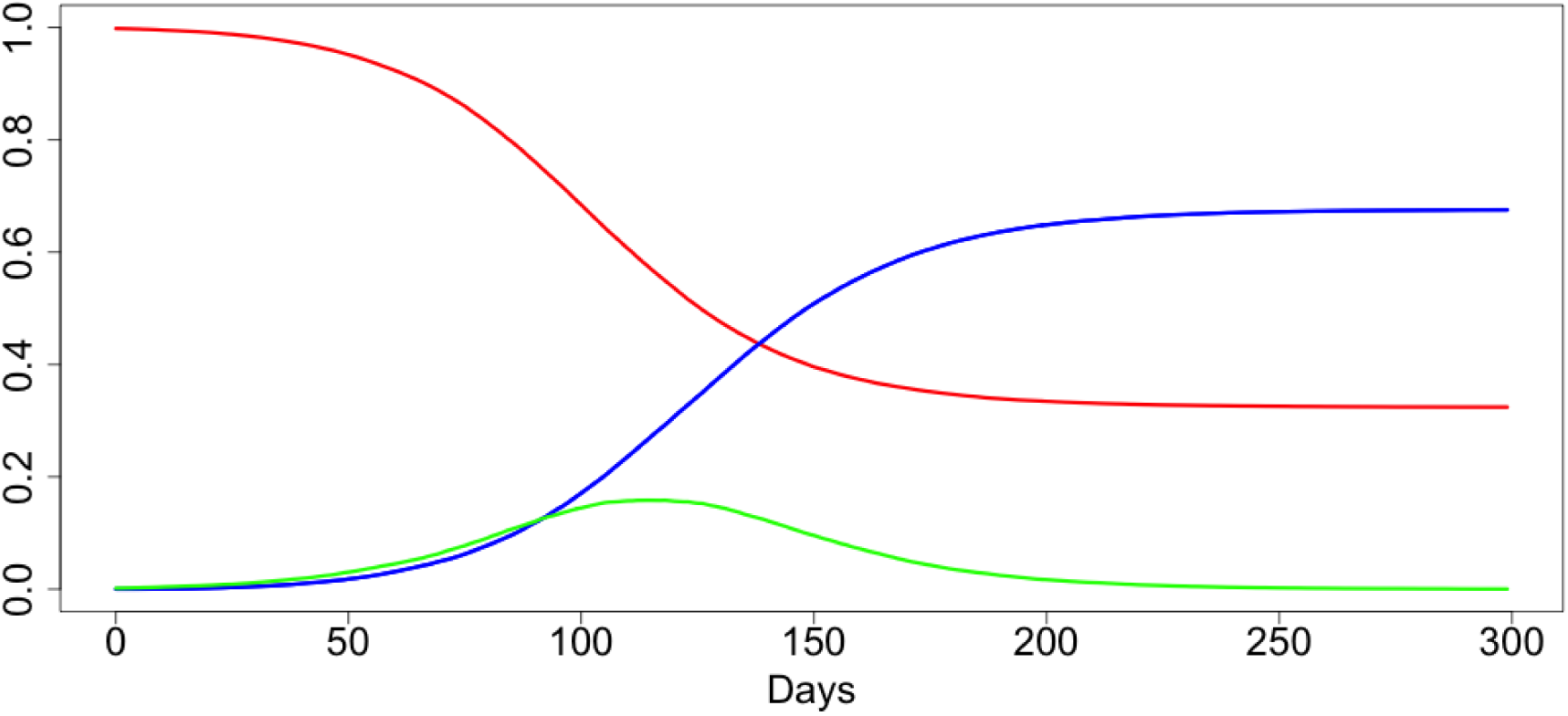
An uninterrupted run of a COVID-19 epidemic in homogeneous conditions

In red colour, in Figure 1 and all plots below we plot values of *S*(*t*)*/N*, the proportion of people non-infected by time *t*. In blue, we plot *R*(*t*)*/N*, the proportion of people recovered from the disease (or dead) and in green we plot our main quantity of interest which is *I*(*t*)*/N*, the proportion of infected people at time *t*. The values of *R*(*t*) do not play any part in modelling and are plotted for information only.

The duration of time while a person with a virus is infected is modelled by Erlang distribution with shape parameter *k* = 3 and mean 21. The simulations are flexible and we can easily change these values and plot similar curves with updated data. Despite the full recovery taking slightly longer, 1*/σ* = 21 days is a good estimation for the period of time when people with severe infection may require intensive care, ventilator etc. Also, we believe that this is a suitable distribution for the period of time when such people may die.

From the value *I*(*t*), we can estimate the distribution of the number of deaths, at time *t*, as follows: first, since on average an infected person is considered as infected for 21 days, we compute *I*(*t*)*/*21. At each particular day, any person can die with the probability which is typical for the chosen population or sub-population. If we consider the whole population, then we apply the mortality rate for the population. However, if *I*(*t*) refers to a particular group only, then we should apply the corresponding coefficient for the group. In any case, the distribution for the number of deaths at time *t* can be roughly considered as binomial with parameters *I*(*t*) and the probability of ‘success’ *rσ*, where *r* is the mortality rate. Different authors disagree on the values of mortality *r* for the COVID-19; see, for example, [10]. UK’s experts believe *r* ≃ 0.009 [3], WHO sets the world-wide mortality rate at 0.034, the authors of [11] believe *r* is very small and could be close to 0.001, an Israeli expert D. Yamin sets *r* = 0.003, see [12].

**Example 1**. *Assume that the epidemic in Birmingham (with population size N* ≃ 1, 086*m) was running uninterrupted according to the scenario depicted in Figure 1. The maximum value of I*(*t*)*/N is* ≃ 0.163 *giving the maximum expected daily death toll of* 0.163 1086000 0.009*/*21 ≃ 75.8 *assuming we use UK experts value r* = 0.009.

Although simple to implement, the estimation in Example 1 is likely to be wrong, as it would be for any heterogeneous city (e.g. London). Critically, as will be discussed in the next section, the maximum death toll is likely to be significantly lower than the above calculations suggest.

Summarizing, the expected daily mortality curve is simply a suitably scaled version of *I*(*t*)*/N* of Figure 1. The same is applied to the curves representing expected number of hospital beds and ventilators. The variability for the number deaths is naturally considerably higher than the variability for the numbers of required hospital beds and ventilators as the latter ones are correlated in view of the fact that each person occupies a bed (requires a ventilator) for a few days in a row but dies only once.

Unlike *I*(*t*), the number of deaths *D*(*t*) at each particular day is (approximately) known and potentially can be used for estimating the first and second derivatives of *I*(*t*)*/N* and hence the stage of the epidemic. It is, however, difficult to do for the following reasons: randomness in the values *D*(*t*), see above, and, more importantly, heterogeneity of large sub-populations, see next section.

One of the main targets of decision-makers for dealing with epidemics like COVID-19 can be referred to as ‘flattening the curve’, where ‘the curve’ is *I*(*t*)*/N* or any of its equivalents and ‘flattening’ roughly means ‘suppressing the maximum’. We shall consider this in the next sections.

## 3 Spatial heterogeneity of the population and heterogeneity of epidemic development in different areas

The purpose of this section is to demonstrate that heterogeneity of epidemic developments for different sub-populations has significant effect on ‘flattening the curve’.

Already when this paper was completed, the authors learned about a preprint [13] which uses SIR modelling to produce somewhat similar conclusions with more specifics for COVID-19 in the UK. However, SIR modelling using numerous age-classes and many equations for each age-class different across different parts of the country resulting in an astronomical number of parameters; this makes any sensitivity analysis to various parameters uncheckable. We believe that a combination of stochastic models like the present one with standard SIR-based models can help in significantly reducing the number of parameters while also adding more flexibility to a hybrid model.

Consider the following situation. Assume that we have a population consisting of *M* sub-populations (groups) *G*_*m*_ with similar demographic and social characteristics and these sub-populations are subject to the same epidemic which has started at slightly different times. Let the sizes of sub-population *G*_*m*_ be *N*_*m*_ with *N*_1_ + … + *N*_*m*_ = *N*. In all cases, the curves *I*_*m*_(*t*)*/N*_*m*_ are shifted in time versions of the curve *I*(*t*)*/N* of Figure 1.

In Figures 2 and 3, *M* = 2 and the second epidemic started 50 days after the first one (incubation period is set to be 0). In Figures 4 and 5, *M* = 10 and each next epidemic cycle has started 7 days after the previous one. In all cases, we evidence the significant ‘flattening the curve’ phenomenon. In Figures 2–5, the green line is used for the resulting curves *I*(*t*)*/N*. The maximal values of *I*(*t*)*/N* in the examples depicted in Figures 2–5 are 0.124, 0.136, 0.134 and 0.132, respectively. This is significantly lower than the value 0.163 for the original curve. In the assumptions of Example 1, these would respectively lead to the maximum expected number of deaths 57.7, 63.3, 62.9 and 62.7.

**Figure 2:**
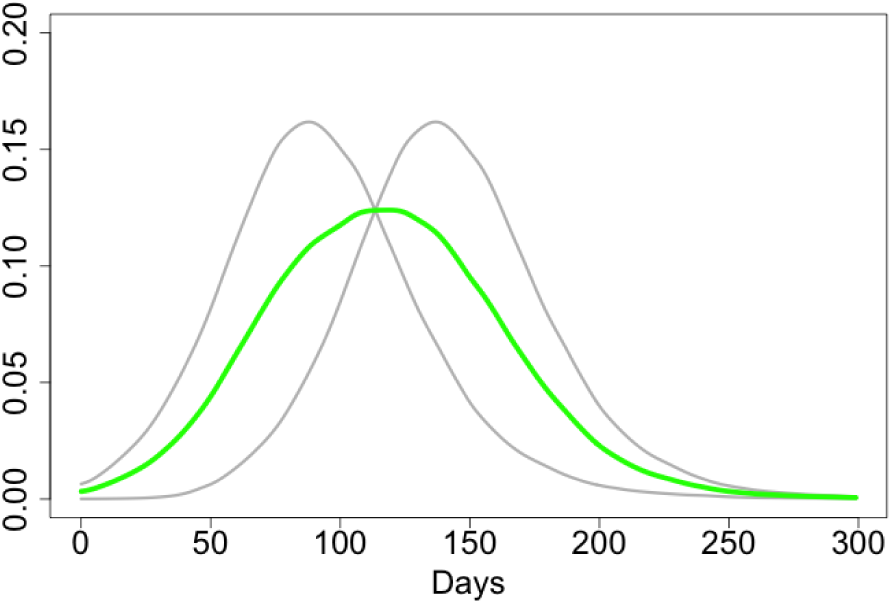
*M* = 2, *N*_1_ = *N*_2_.

**Figure 3:**
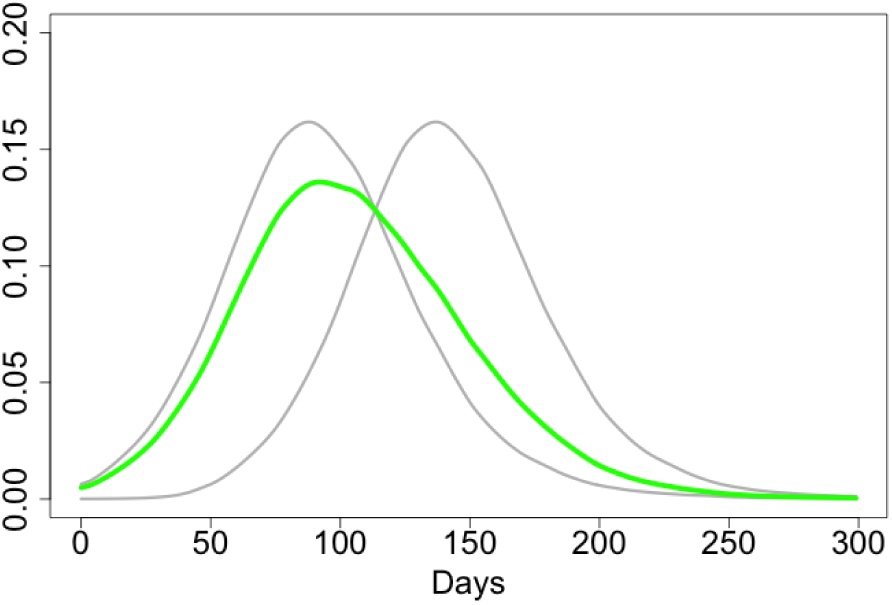
*M* = 2, *N*_1_ = 3*N*_2_

**Figure 4:**
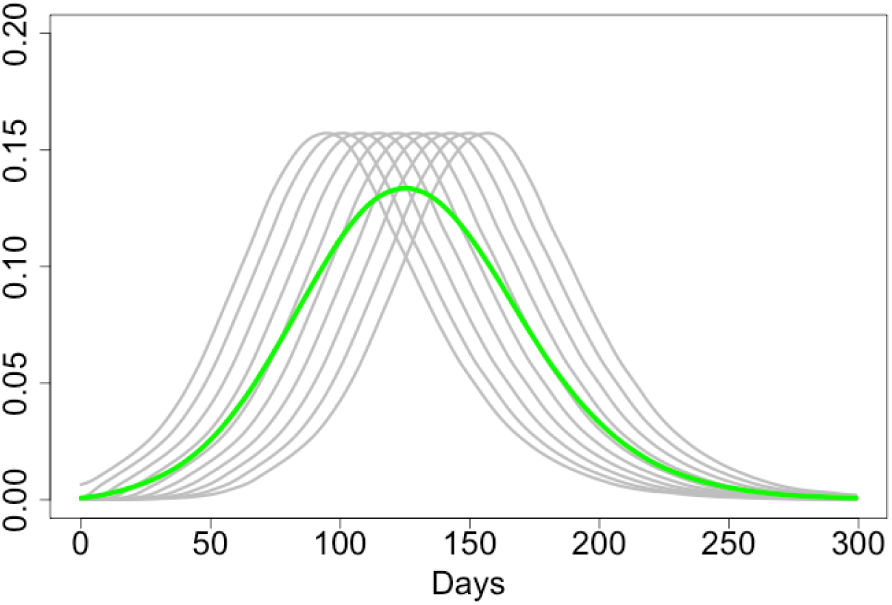
*M* = 10, 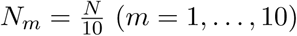

**Figure 5:**
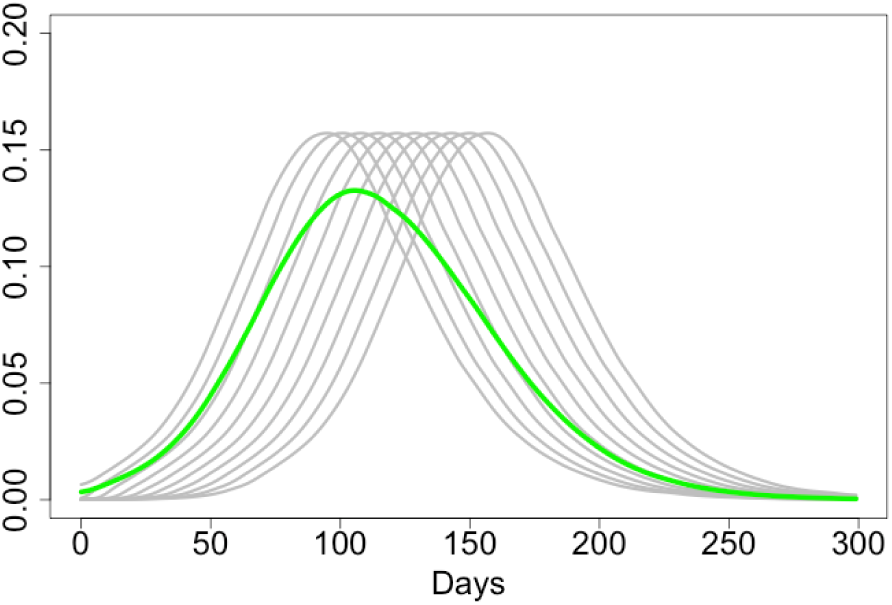
*M* = 10, 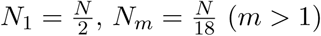

Extra heterogeneity of different epidemics caused by the social and demographic heterogeneity of the sub-populations would further ‘flatten the curve’. The following conclusions are in line with what the Israeli expert D. Yamin states [12] and agree with the main conclusions of the paper [14] which is rather critical towards standard epidemiological models.

**Conclusions**. *(a) Isolation of sub-populations at initial stages of an epidemic is very important for preserving heterogeneity of the epidemic and ‘flattenning the curve’; this flattening can be very significant. (b) Epidemiological models based on the assumption of a homogeneous population but applied for populations consisting of heterogeneous sub-populations may give completely misleading results*.

## 4 An epidemic with intervention

In this section, we assume that we make an intervention to the epidemic by introducing an isolation at a certain stage. Moreover, we assume that isolation may be different for 2 different groups. We define a special group *G* of more vulnerable people consisting, for example, of all people aged 70+. We define *α* = *n/N*, where *N* is the total population size and *n* is the size of this special group *G*.

Let *t*_0_ be the moment of time when the isolation occurs. It is natural to define *t*_0_ from the condition *S*(*t*_0_)*/N* = *x*, where, for example, *x* = 0.9. For *t < t*_0_, the virus has been transmitted to people uniformly so that, conditionally a virus is transmitted, the probability that it reaches a person from group *G* is *α*. For *t ≥ t*_0_, the virus has been transmitted to people in such a way that, conditionally a virus is transmitted, the probability that it reaches a person from *G* is *p* = *cα* with 0 *< c ≤* 1. Moreover, at time *t*_0_ the value of *R*_0_ may change to 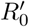 in view of self-isolation. The parameters in this model are:

- *α* = *n/N* : relative size of the group *G*;
- *x ∈* (0, 1) defines the start of the isolation strategy;
- *c* = *p/α* defines the strength of isolation of the group *G*;
- *R*_0_ (initial);
- 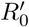: the reproductive number after intervention for *t*≥ *t*_0_; it defines the strength of the overall isolation.

We have run a large number of scenarios coded using a combination of R and Julia [15]; the code is provided in Appendix. In Figures 6–11, we illustrate a few of these scenarios. In all these scenarios, we have chosen *R*_0_ = 2.5 and *α* = 0.2. The results are robust towards values of these parameters (subject to faster or slower rate of the epidemic in dependence on *R*_0_). In Section 5 we use *α* = 0.132 to illustrate some specific results; *α* = 0.2 can be considered either as a generic value or, in the contents of Section 5, as the relative size of the group of vulnerable people which is larger than the group of people aged 70+. We would like to emphasise that there is still a lot of uncertainty on who is vulnerable. It is very possible that the long term effect of the virus might cause many extra morbidities in the coming years coming from severe cases who recover. It is also possible that the virus will mutate and the new strain might affect younger populations more than the current strain. These, and many more possibilities, have a non-negligible probability of occurring, and they have devastating effects. In future models we will try to address these possibilities.

**Figure 6:**
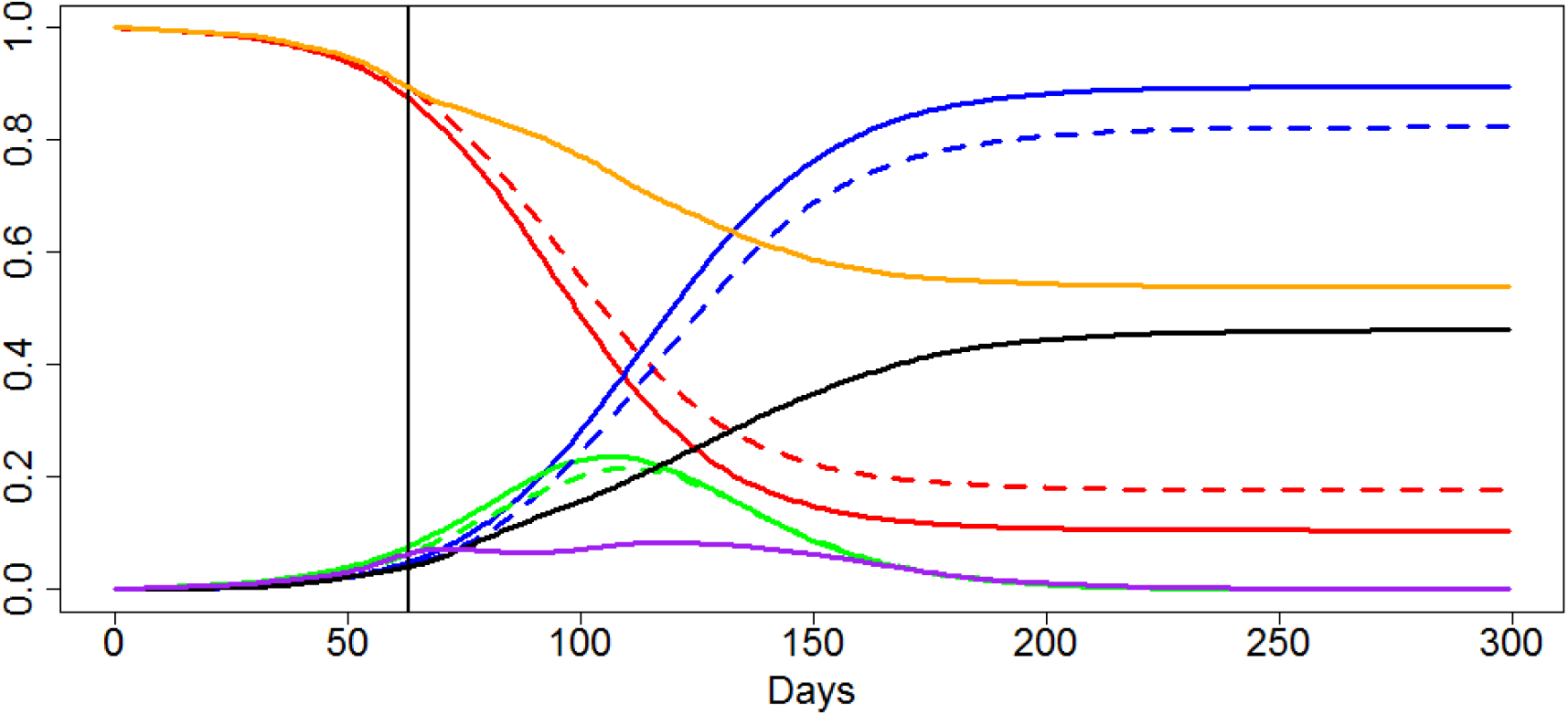
*x* = 0.9, 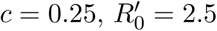

To estimate the number of COVID cases at a given time *t* is difficult [4]. This implies that at the time of making an intervention only very rough guesses about the value of *x*, which is crucial for the future development of the epidemic, can be made.

We distinguish different strengths of isolation by values of parameters *c* and 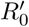:

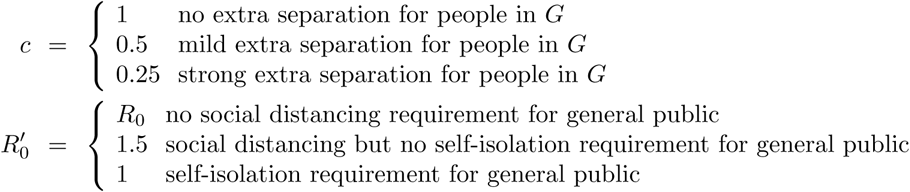

The values *c* = 0.5 and *c* = 0.25 mean that under the condition that a virus is infecting a new person, the probabilities that this new person belongs to *G* are 1*/*3 and 1*/*5 respectively.

Measuring the level of compliance in the population and converting this to simple epidemiological measures *c* and 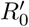 is hugely complex problem which is beyond the scope of this paper.

The lines and respective colours in Figures 6–11 are as follows.

- Solid blue line: *R*(*t*)*/N*, the proportion of people recovered from the disease (or dead) in case of no intervention, as in Figure 1
- Dashed blue line: *R*(*t*)*/N* in case of intervention
- Solid black line: *R*_*G*_(*t*)*/n* the proportion of people from group *G* recovered from the disease (or dead) in case of intervention
- Solid red line: *S*(*t*)*/N*, the proportion of people non-infected by time *t* in case of no intervention, as in Figure 1
- Solid orange line: *S*_*G*_(*t*)*/n*, the proportion of people susceptible to the virus from group *G* non-infected by time *t* in case of intervention
- Solid green line: *I*(*t*)*/N*, the proportion of infected people at time *t* with no intervention, as in Figure 1
- Dashed green line: *I*(*t*)*/N*, the proportion of infected people at time *t* with intervention
- Solid purple line: *I*_*G*_(*t*)*/n*, the proportion of infected people from group *G* at time *t* with intervention

Considerably important, in view of the discussions of the next section, is Figure 6. In this figure, for the initial data of Figure 1, we strongly separate group *G* at the time when 10% of the population is infected. We make no call to the general public for social distancing. In this scenario, the intervention does not considerably change the proportion of infected in the total population but it significantly ‘flattens the curve’ for the group *G*: compare the purple and green lines. The maximum of *I*_*G*_(*t*)*/n* is 0.086 which is 2.5 times lower than 0.216, the maximum of *I*(*t*)*/N*. Note also the fact that the curve *I*_*G*_(*t*)*/n* is rather flat for long time, during the main stage of development of the epidemic. Just after the peak of the epidemic in the whole population, *I*_*G*_(*t*)*/n* peaks; it is caused by the presence of very large number of infected people from the general population.

The scenario which led to Figure 6 is important and hence it has been re-run for a few variations of the model. Figure 7 shows the results of the simulations where we have removed the incubation period. The maximum of *I*_*G*_(*t*)*/n* is now 0.075 which is more than twice lower than 1.63, the maximum of *I*(*t*)*/N*. Note that in the scenario with no incubation period the whole epidemic is milder as the virus lives longer. In Figure 8 we use similar scenario as for Figure 6 but we separate group *G* only mildly at the time when 10% of the population is infected. We make no call to general public for social distancing. The curve *I*_*G*_(*t*)*/n* is ‘flattened’ for the group *G* but in a considerably smaller degree than in the case of strong separation of *G*.

**Figure 7:**
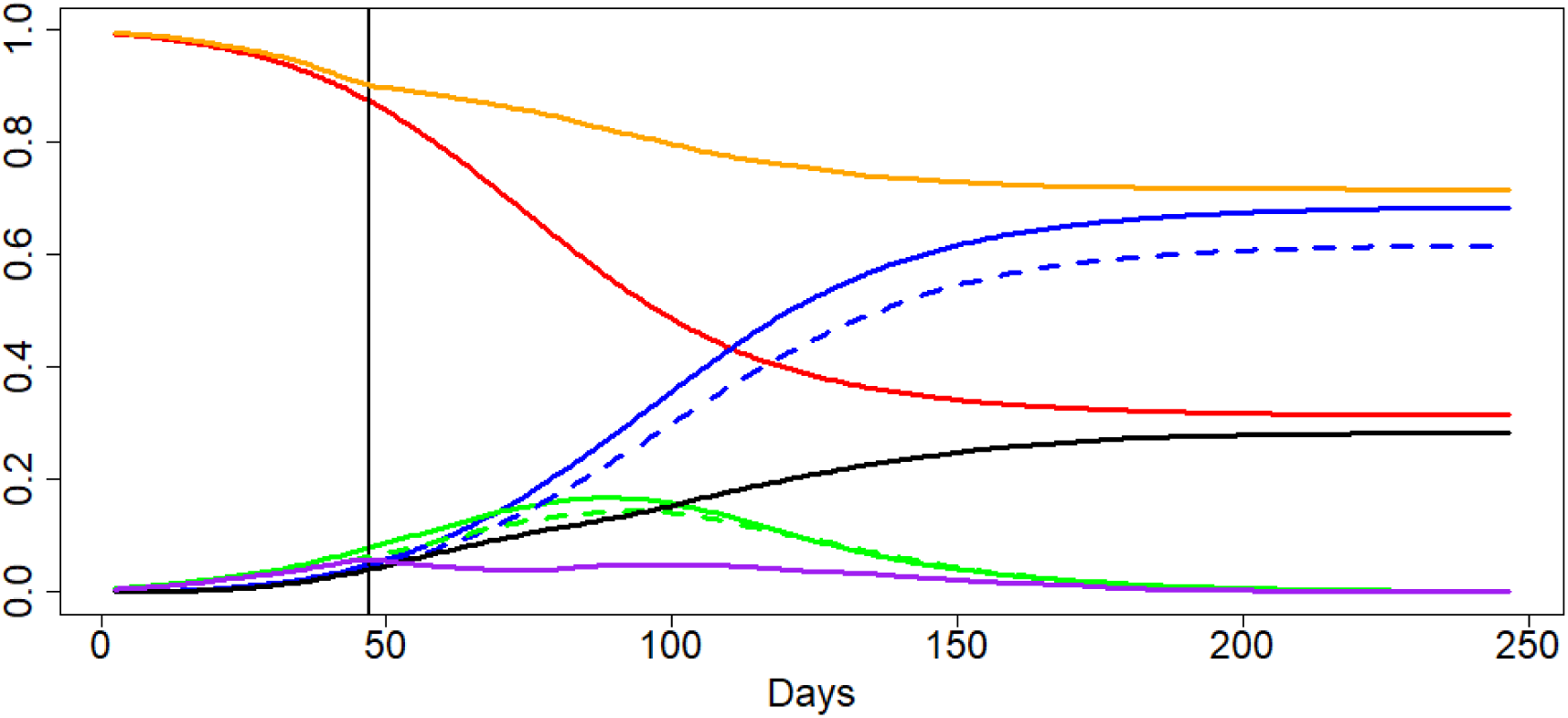
*x* = 0.9, *c* = 0.25, 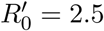, no incubation period

**Figure 8:**
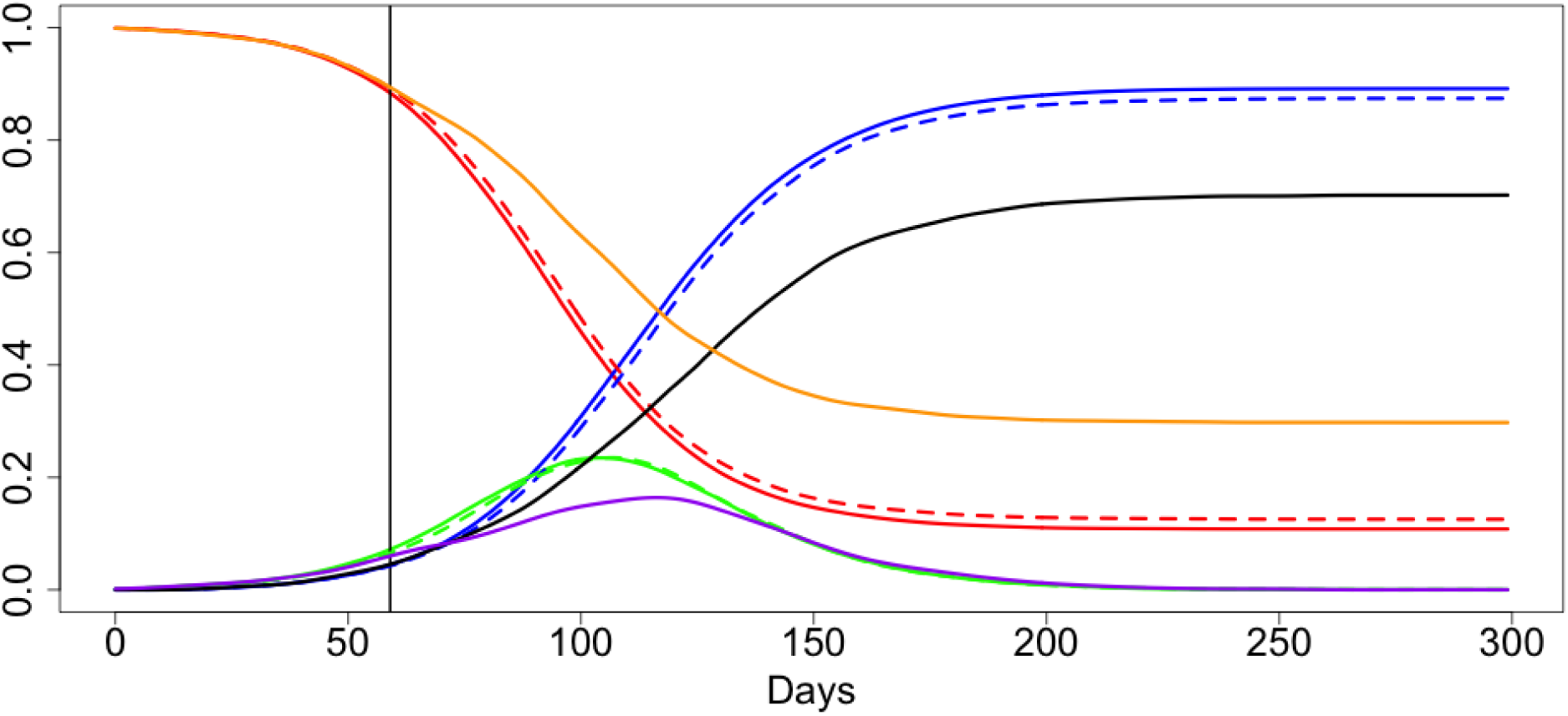
*x* = 0.9, *c* = 0.5, 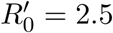

Figures 9 and 10 illustrate the situation with mild and strong separation of people from *G* complemented with introduction of the social distancing for general public. Interestingly enough, the effect of social distancing for general public gives less benefit than even mild isolation of people from group *G*.

**Figure 9:**
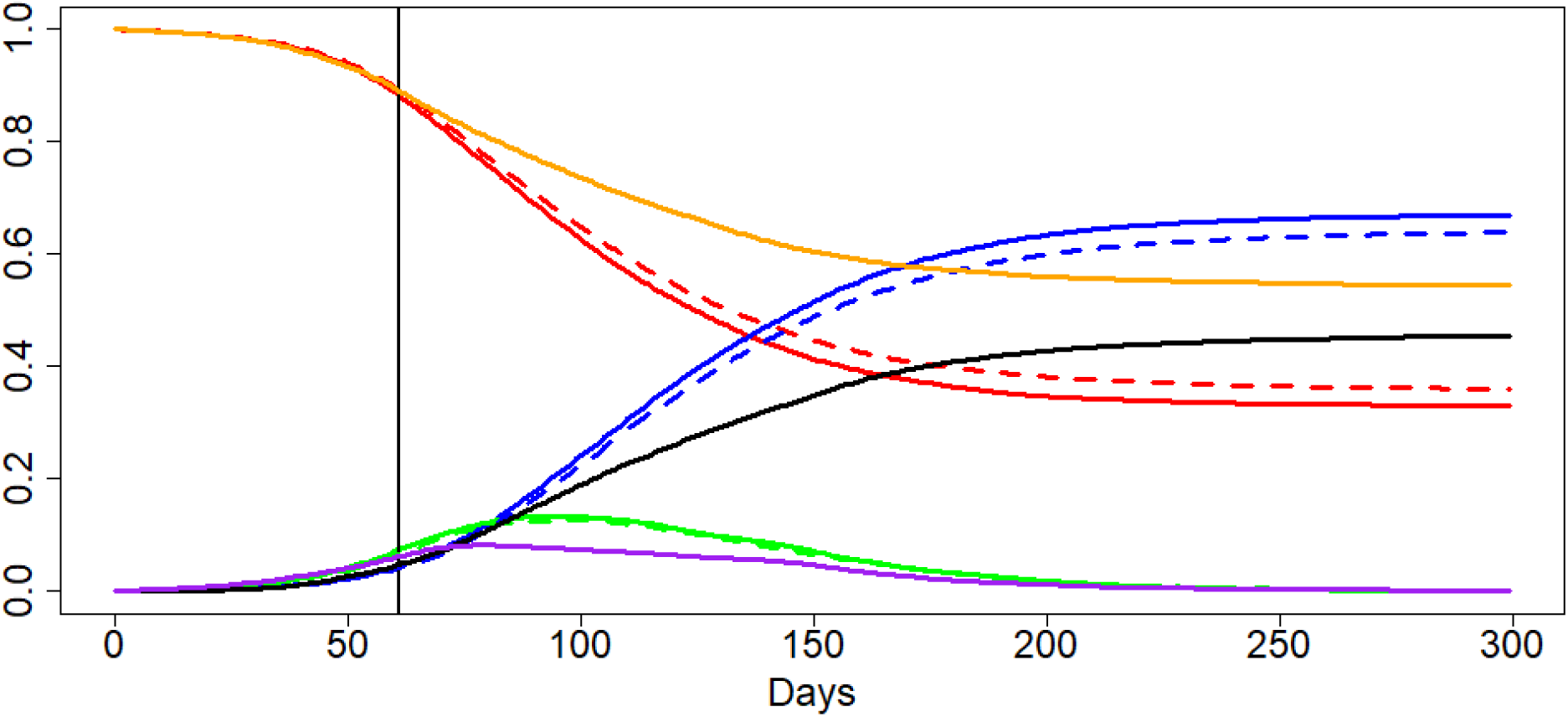
*x* = 0.9, *c* = 0.5, 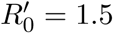

**Figure 10:**
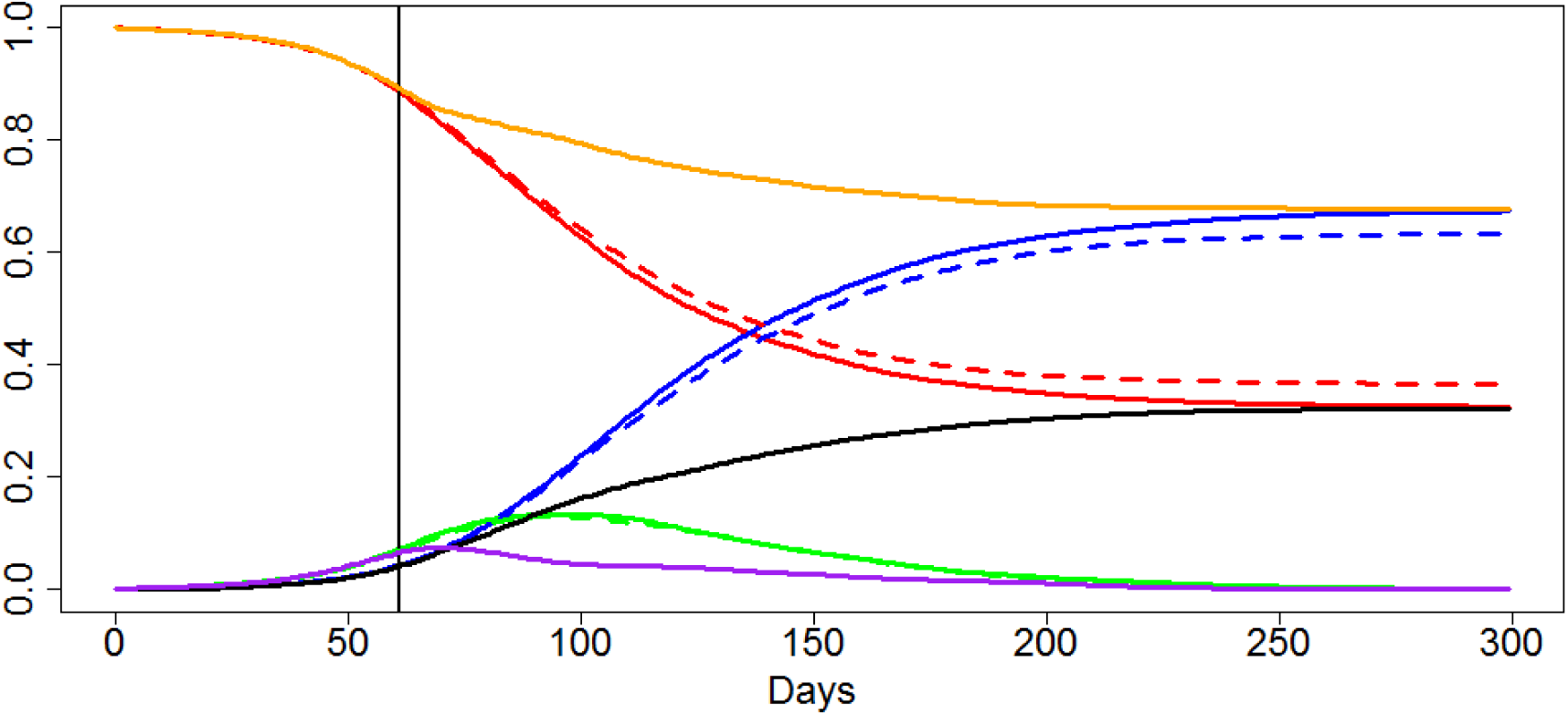
*x* = 0.9, *c* = 0.25, 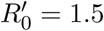

Figure 11 illustrates the scenario with no call to the general public for social distancing but with strong separation of the people from *G* at at a later stage of epidemic, when 20% of the population is infected. The effect is similar to the one observed in Figures 6 and 7 except for the fact that the call for isolating the group *G* came slightly late.

**Figure 11:**
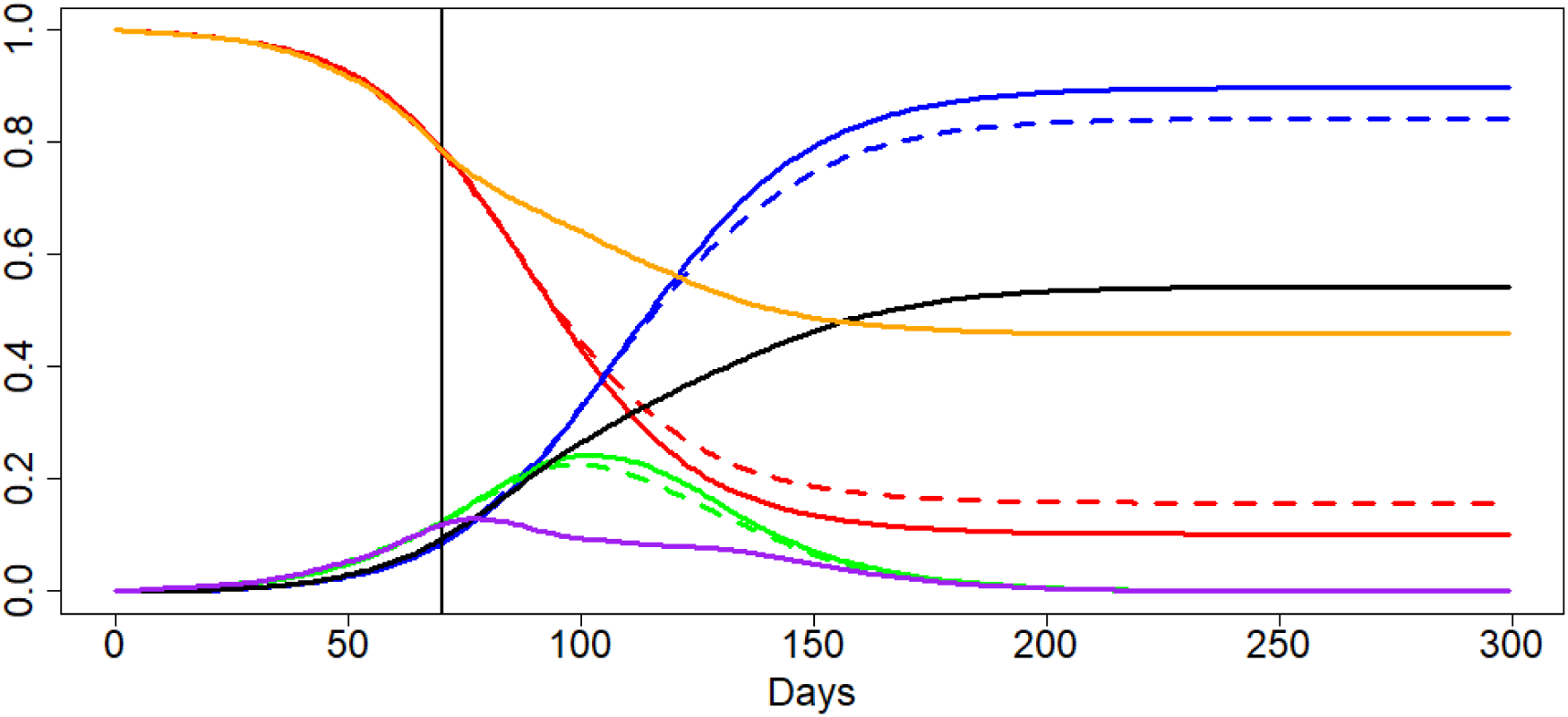
*x* = 0.8, *c* = 0.25, 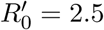

**Conclusion**. *By considering a number of scenarios we have observed an extreme sensitivity of the negative consequences of the epidemic to the degree of separation of vulnerable people. Sensitivity to the parameter measuring the degree of self-isolation for the whole population is less apparent although it is much costlier*.

This conclusion is very much in line with recommendations of leading German epidemiologists [16].

## 5 Consequences for the mortality and impact on NHS

Results of the type presented in Figures 6–11 can be translated into the language of the expected number of death and expected number of beds required. To do this we extend the observations we made at the end of Section 2 concerning translation of the curve *I*(*t*)*/N* into the curves for the expected number of death *ED*(*t*) and expected number of hospital beds in the UK. We use the common split of the UK population into following age groups:

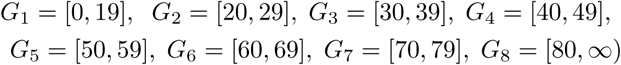

and corresponding numbers *N*_*m*_ (*m* = 1, …, 8; in millions) taken from [18]

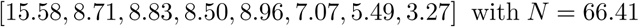

The death probabilities *p*_*m*_ are given from Table 1 in [3] and replicated many times by the BBC and other news agencies are:

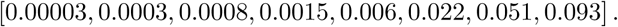

Unfortunately, these numbers do not match the other key number given in [3]: the UK average mortality rate which is estimated to be about 0.9%. As we feel the value of the UK average mortality rate is more important, we have multiplied all probabilities above by 0.732 to get the average mortality rate to be 0.9%.

Defining the group *G* as a union of groups *G*_7_ and *G*_8_, we have *n* = 5.49 + 3.27 = 8.76m and *α* = 8.76*/*66.41 ≃ 0.132. We then compute the mortality rate in the group *G* and for the rest of population by

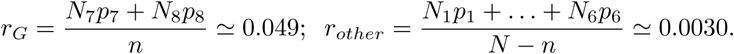

For estimating the average number of death in the group *G* and for the rest of population we can the use the formulas

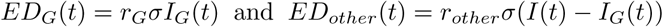

respectively. The average numbers of hospital beds required for two different groups are proportional to these numbers.

We have run a series of scenarios for an UK epidemic without taking into account spatial and social heterogeneity of the society assuming that we would have separated (mildly and strongly) the group *G* of 70+ people at the time *t*_0_ when 10% of the population were infected with no call to general public for social distancing. We marked *t*_0_ as March 23, 2020.

The two scenarios (with *c* = 0.5 and *c* = 0.25) we have used for illustrating this technique are different from the scenarios used for plotting Figures 6 and 7 only by the value of *α*. For Figures 6 and 7, we have chosen *α* = 0.2 but for the group *G* of 70+ old people in the UK we have *α* = 0.13.

In Figures 12 and 13 we plot (using the same colours as in Figures 6 and 7) the following curves: *I*(*t*)*/N* (solid green, no intervention on March 23), *I*(*t*)*/N* (dashed green, intervention on March 23) and *I*_*G*_(*t*)*/n* (purple, intervention on March 23).

**Figure 12:**
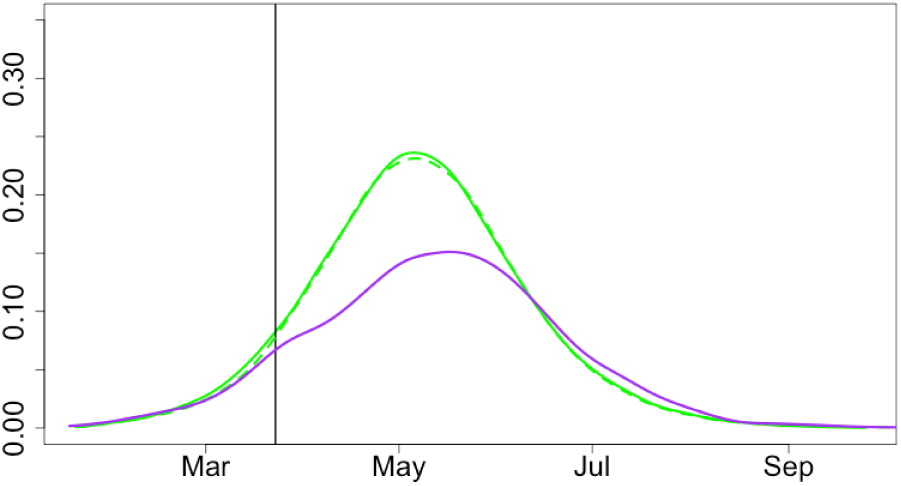
*c* = 0.5

**Figure 13:**
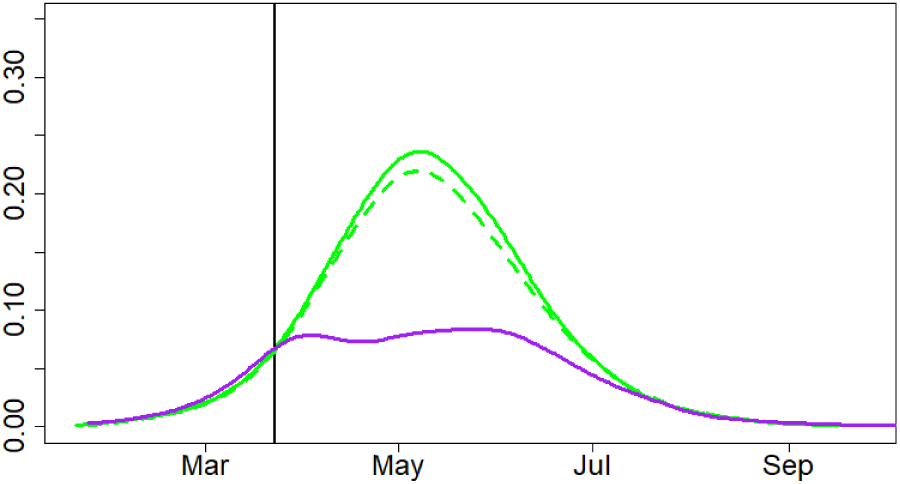
*c* = 0.25

In Figures 14 and 15 we plot the curves for the estimated average number of deaths: *ED*(*t*) (solid green, no intervention on March 23), *ED*_*G*_(*t*) (purple, intervention on March 23), *ED*_*other*_(*t*) (dashed green, intervention on March 23) and combined *E*[*D*_*G*_(*t*) + *D*_*other*_(*t*)] (black, intervention on March 23). We can deduce from Figure 15 (taking into account the extra factor of hospital beds availability) that strong separation of 70+ old people alone would have reduced the expected number of death by at least the factor of 2. Another feature of the scenario with *c* = 0.25 is a roughly 50/50 split between the number of deaths in the 70+ and 70− groups.

**Figure 14:**
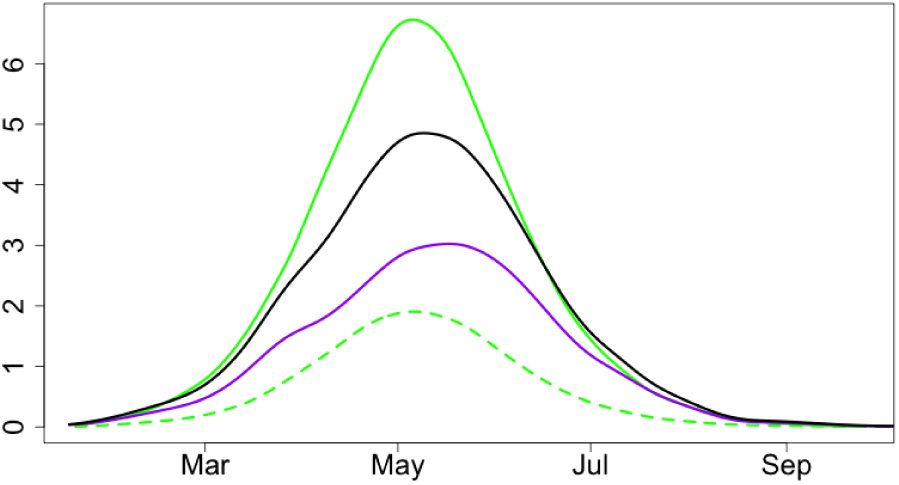
*c* = 0.5

**Figure 15:**
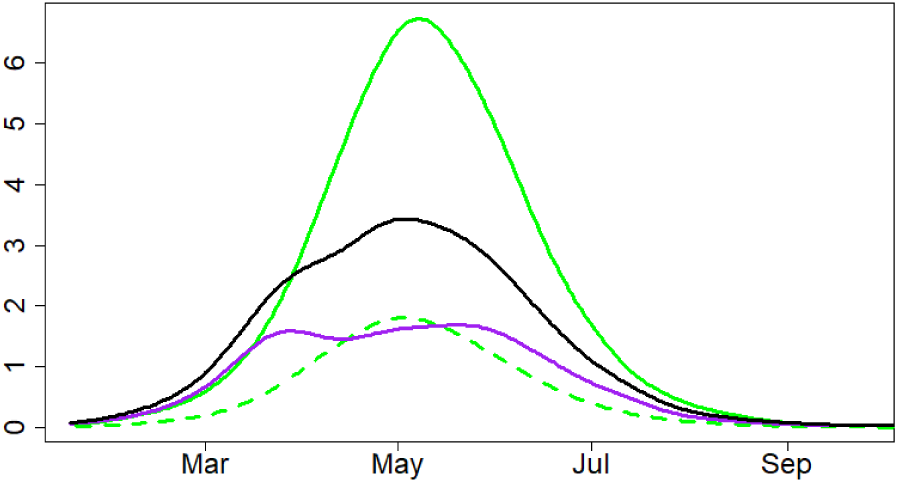
*c* = 0.25

The y-axis in Figures 14 and 15, after multiplication by 140, can be roughly interpreted as hundreds in the London epidemic assuming *R*_0_ = 2.5, *x* = 0.9 on March 23 and homogeneity of the epidemic (as there is about 9m people in London out of 64.1m). As mentioned in Section 3, deaths numbers in other regions of the UK should be expected to be lower.

We then have run through the scenarios with partial lockdown on March 23 reducing *R*_0_ = 2.5 to 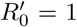 and with a return, 30 days later, to *R*_0_ = 2.5 with *c* = 0.5 and *c* = 0.25. Results are plotted in Figures 16-19. The style of Figure is exactly the same as for Figures 12-15. If the value *x* (proportion of non-infected people on March 23) happens to be larger than 0.9 then the second wave of epidemic should be expected to be (perhaps, considerably) larger. If *x <* 0.9 then the second wave will be less pronounced.

**Figure 16:**
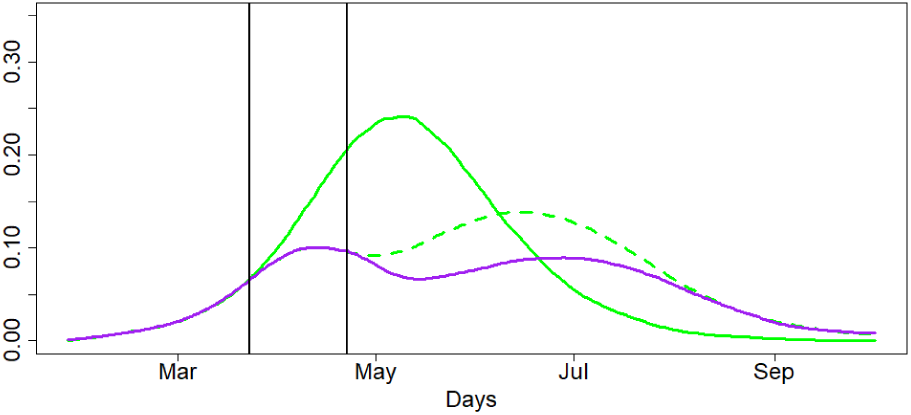
*c* = 0.5

All these figures are given for illustration only without any claim on accurate predictions as there is an uncertainty of the outputs towards the choice of several parameters describing the virus characteristics but the sensitivity of the model towards the choice of these parameters is not yet adequately assessed. Figures 20-21 illustrate sensitivity of the scenario of Figures 16-17 with respect to the choice of *k*, the shape parameter of the Erlang distribution defined in Section 2. In these figures, we plot 100 trajectories similar to the trajectories of Figures 16-17 with the values of *k* taken at random in the interval [2, 4].

**Figure 17:**
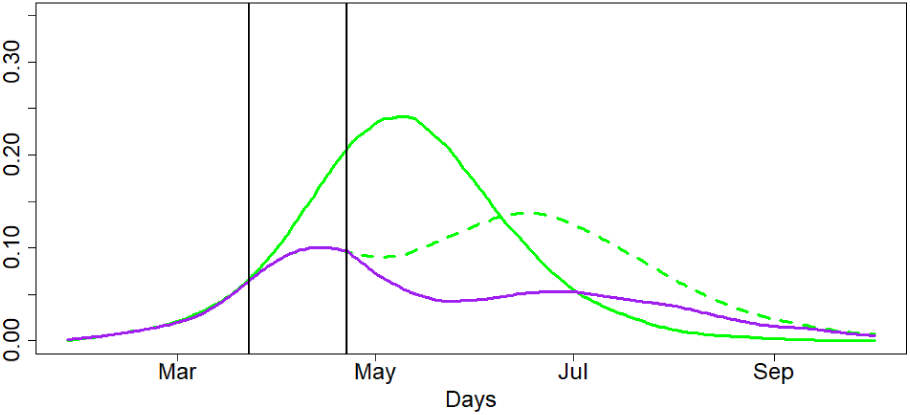
*c* = 0.25

**Figure 18:**
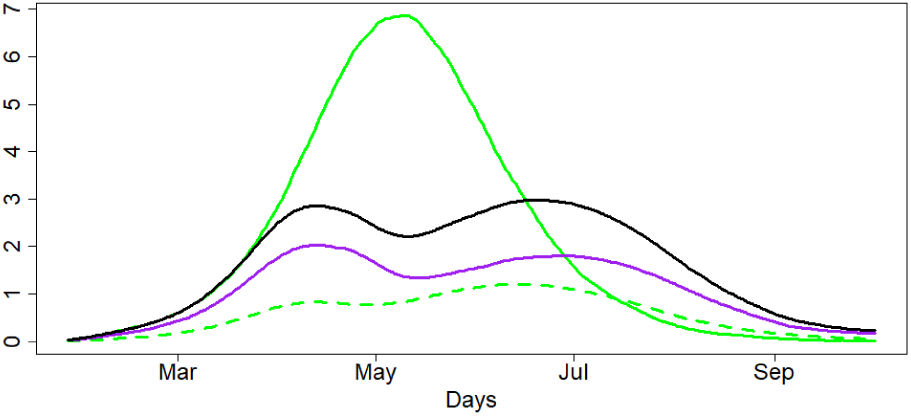
*c* = 0.5

**Figure 19:**
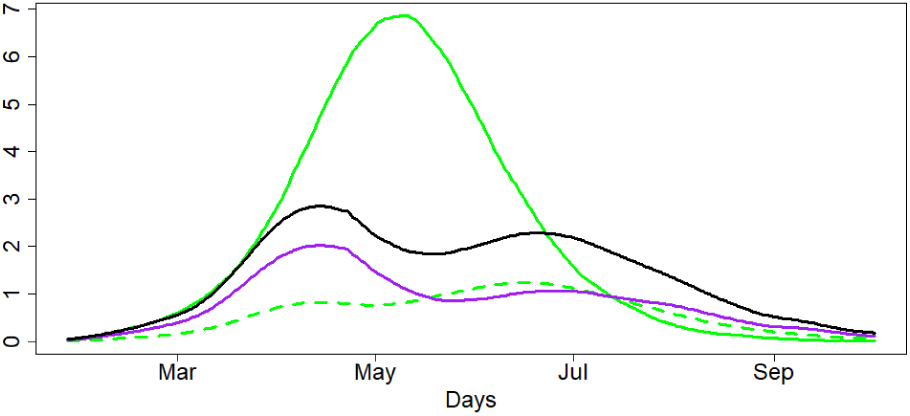
*c* = 0.25

**Figure 20:**
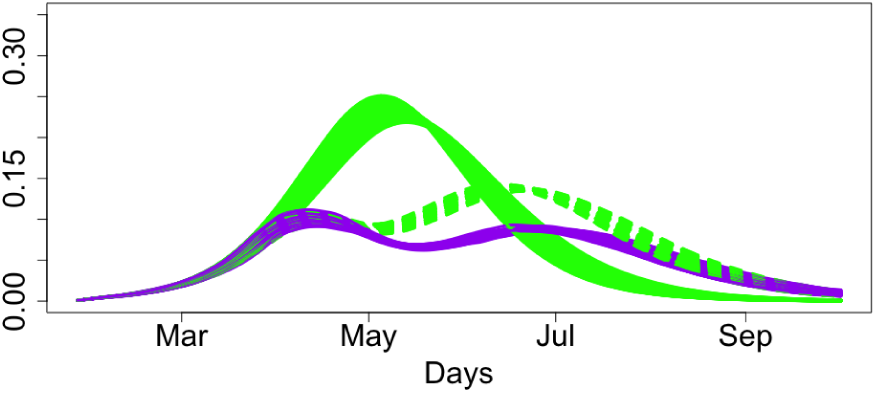
*c* = 0.5, ∈ [2; 4]

**Figure 21:**
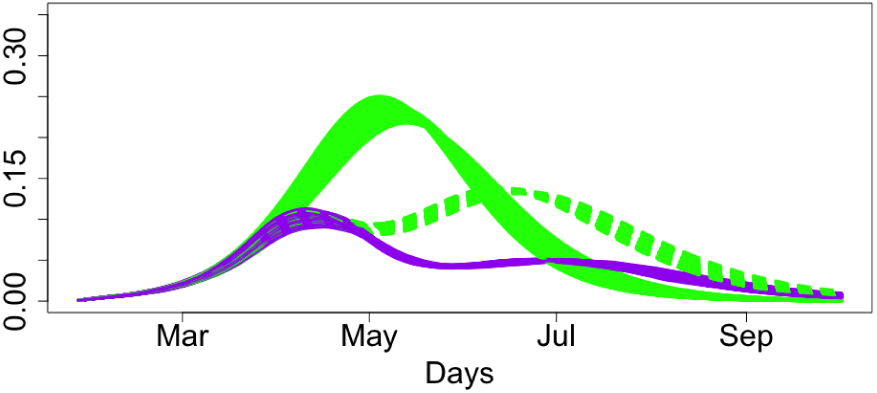
*c* = 0.25, ∈ [2; 4]

**Conclusion**. *The outputs of the model developed above can be translated into the language of the expected number of death due to the epidemic. This language amplifies the findings of the previous section stating that isolation of a relatively small percentage of population may hugely reduce the death toll of the epidemic*.

### Some of the next steps and open problems

- Run many different scenarios to get better understanding of the current situation with the epidemic and what can be done to effectively control it.
- Continue calibrating the model against existing epidemiological models and new data as it emerges. By using methods of stochastic global optimization [18, 19], teach the model to learn from the data emerging daily, for adapting values of parameters describing the virus and hence the course of the epidemic.
- Incorporate into the model algorithms for quickest on-line detection of changes [20, 21] to learn about temporal and spatial heterogeneity of the development of the epidemic.
- Understanding the reproductive number *R*_0_ in dependence on the population density in local areas and hence the mixing distribution for *R*_0_ needed to combine sub-populations.
- Understanding *R*_0_ and other virus parameters as random variables due to mutations.
- Understanding the risk as a function depending on different factors such as age and social groups. Use of OR models for studying NHS capacity issues under different scenarios and correlated to the percentage of health providers getting infected by the virus.
- Properly quantify uncertainty for model predictions.
- Full-scale sensitivity analysis to different parameters.
- With better understanding of the role of parameters, formulate inverse problems like finding the stage of an epidemic by its early development.
- Use game theory [22] to produce tools for optimal decision making with respect to slowing down the process of epidemic by enforcing spatial isolation and isolation of different sub-populations.
- Develop a more sophisticated model combining stochastic differential calculus [23], fractional diffusion [24] and elements of direct simulation (the models of fractional diffusion will be used to understand the effects of slowing down of the epidemic).
- Understand and quantify the differences in epidemic developments across different countries such as UK, USA, Italy and Spain.
- Studying long-term effects of coronovirus and future mortality from Covid-19.
- Studying mortality from other diseases due to the capacity/fragility of the health system during the epidemic.
- Modelling epidemic control strategies based on testing, tracing and isolation.

## Data Availability

The work is based on simulations only and described in the paper.

## Acknowledgement

We are grateful to several colleagues in Univ. Cambridge and Univ. Warwick for useful comments on drafts of this paper and to M. Hairer (ICL) and J. Ball (Univ. Nottingham) for valuable comments and discussions.

## Appendix: Julia/R code used for computing the scenarios of Section 4

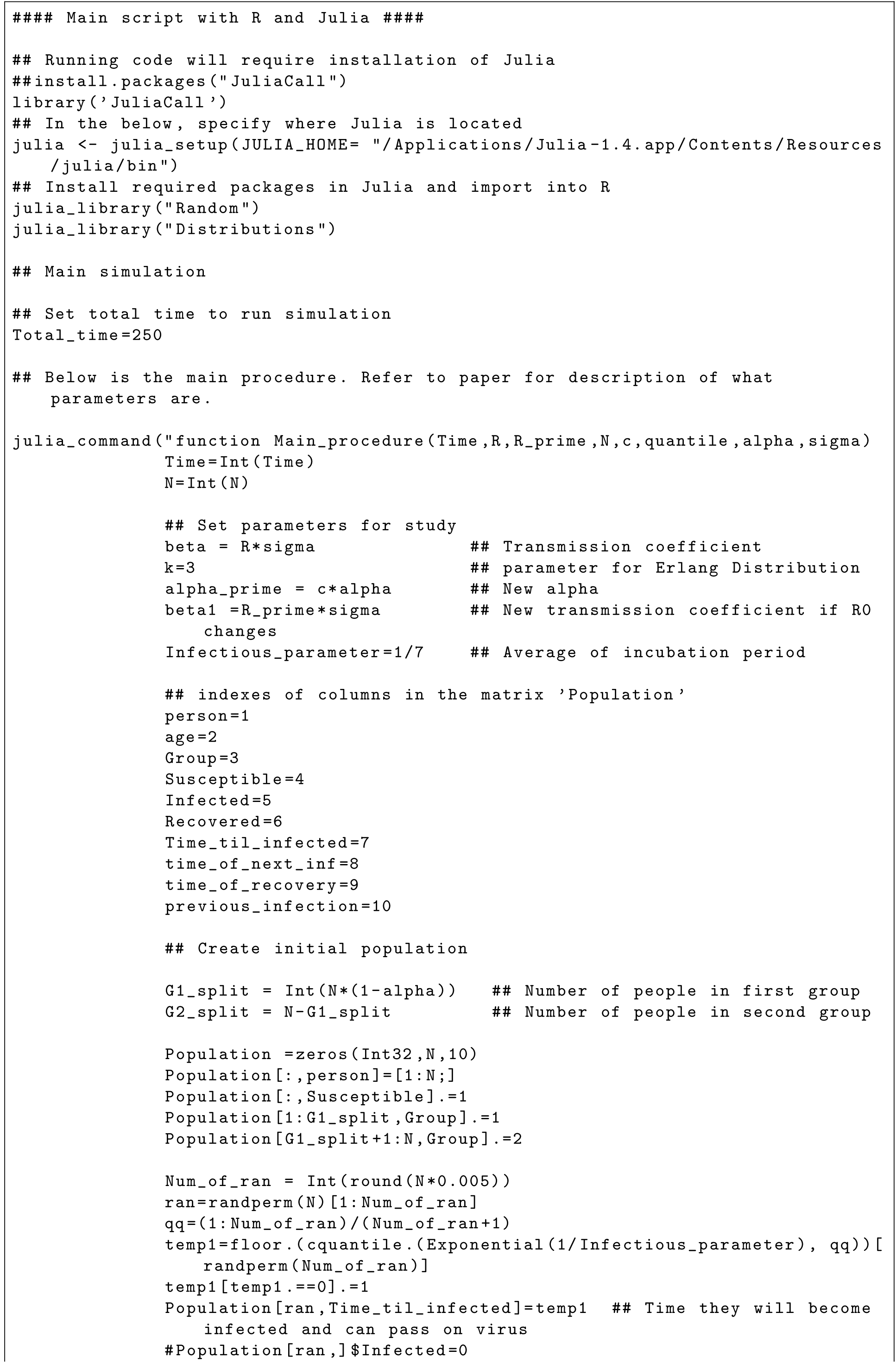
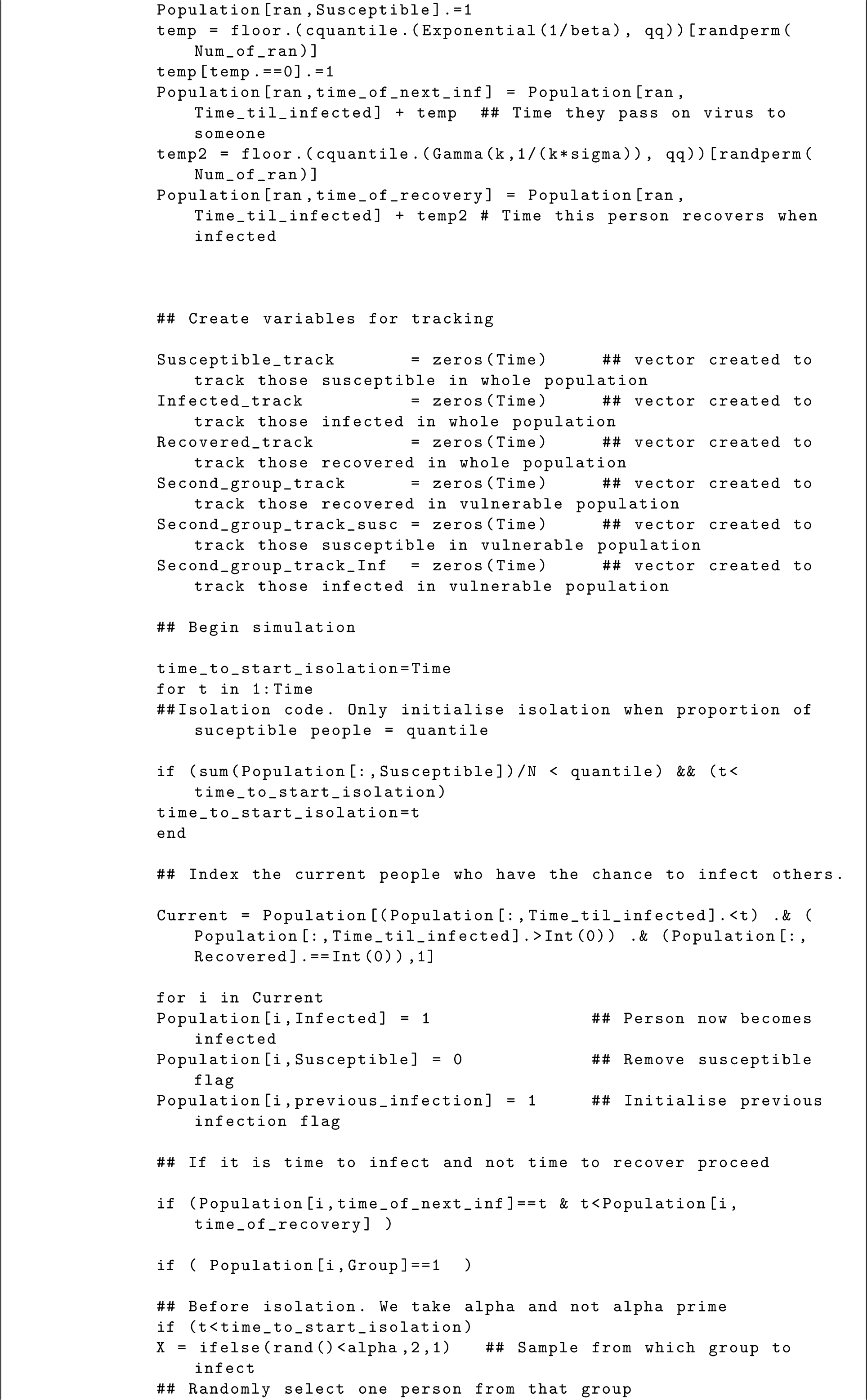
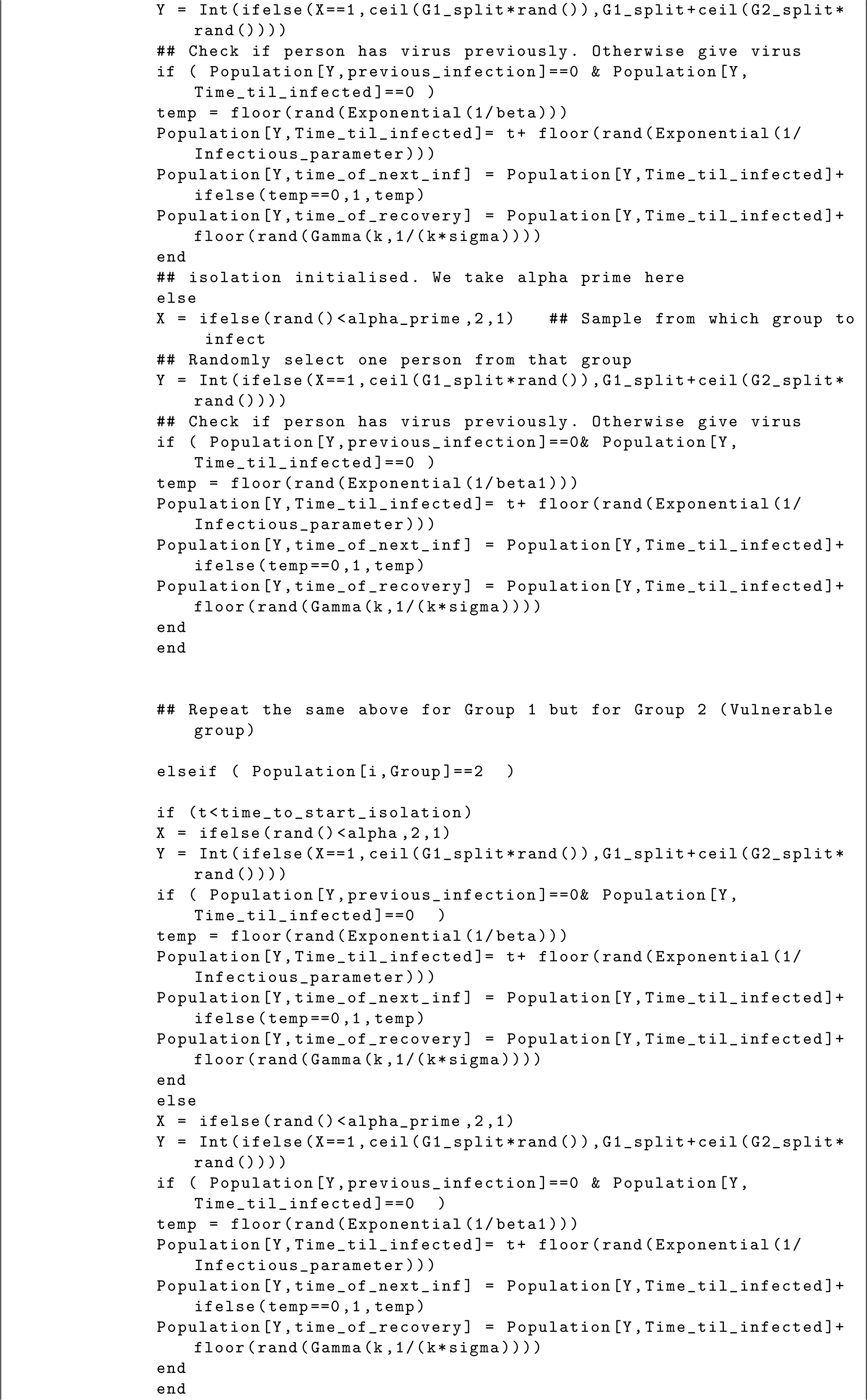
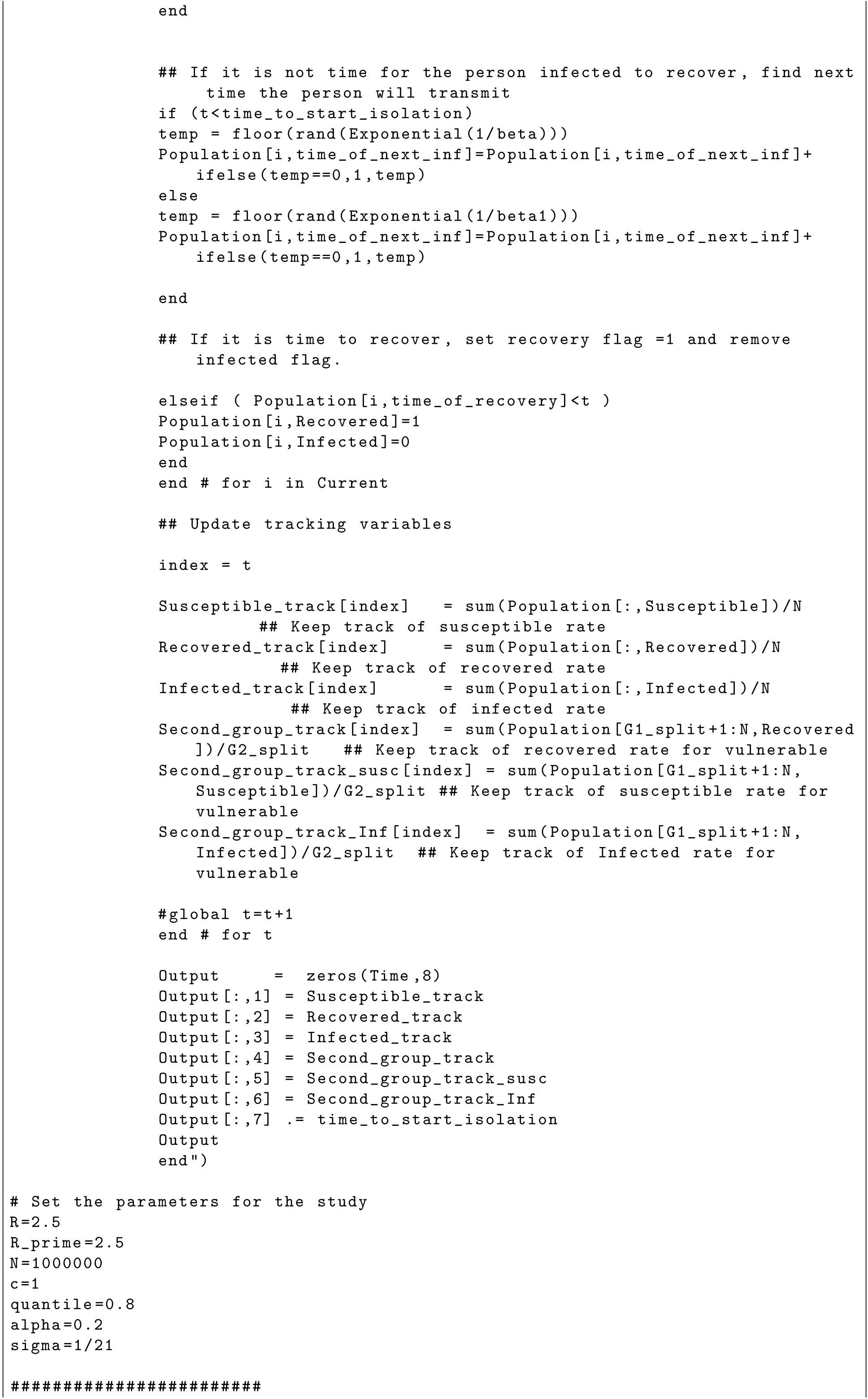
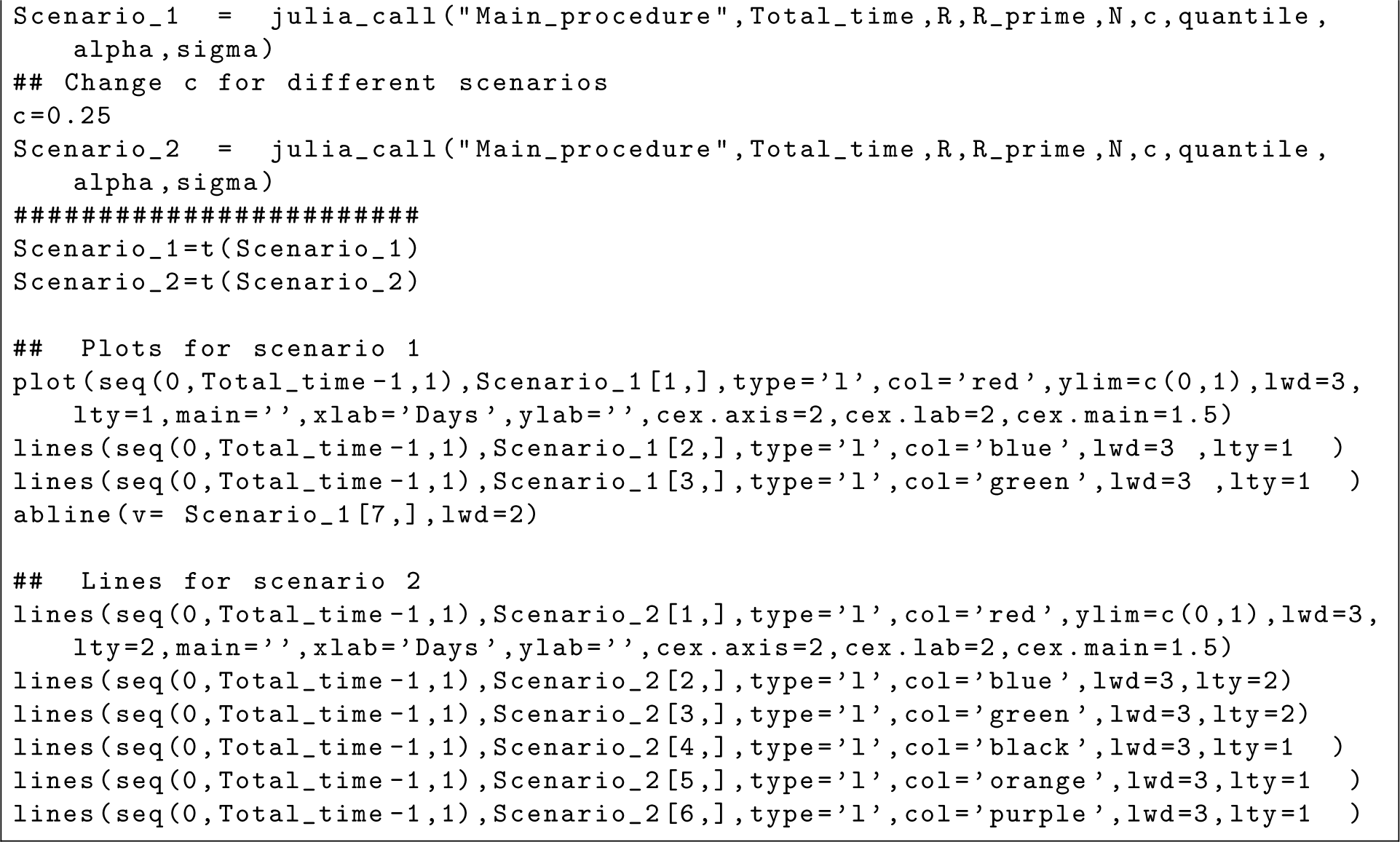

